# Asymptomatic and presymptomatic transmission of SARS-CoV-2: A systematic review

**DOI:** 10.1101/2020.06.11.20129072

**Authors:** Christina Savvides, Robert Siegel

**Affiliations:** Department of Biomedical Informatics, Stanford University, Stanford, California, USA; Department of Microbiology and Immunology, Stanford University, Stanford, California, USA

**Keywords:** presymptomatic transmission, asymptomatic transmission, RT-PCR, viral load, natural history of infection, SARS-CoV-2, systematic review, viral kinetics, serial interval

## Abstract

**Background and Purpose:** Many of the statutes comprising the shelter-in-place and phased-reopening orders are centered around minimizing asymptomatic and presymptomatic transmission. Assumptions about the presence and relative importance of asymptomatic and presymptomatic transmission are based on case reports, the failing of quarantine measures aimed at sequestering ill patients, viral dynamic studies suggesting SARS-CoV-2 production peaks before symptoms appear, and modeling evidence that calculates serial interval between successive generations of infection. In aggregate, these data offer compelling evidence of asymptomatic and presymptomatic transmission, but individually these studies have notable shortcomings that undermine their conclusions. The purpose of this review is to discuss the literature of asymptomatic and presymptomatic transmission, highlight limitations of recent studies, and propose experiments that, if conducted, would provide a more definitive analysis of the relative role of asymptomatic and presymptomatic transmission in the ongoing SARS-CoV-2 pandemic.

**Methods:** We conducted a systematic review of literature on PubMed using search filters that relate to asymptomatic and presymptomatic transmission as well as serial interval and viral dynamics. We focused on studies that provided primary clinical data.

**Results:** 34 studies were eligible for inclusion in this systematic review: 11 case reports pertaining to asymptomatic transmission, 9 viral kinetic studies, 13 serial interval studies, and 1 study with viral kinetics and serial interval.

**Conclusion:** Different approaches to determining the presence and prevalence of asymptomatic and presymptomatic SARS-CoV-2 transmission have notable shortcomings, which were highlighted in this review and limit our ability to draw definitive conclusions. Conducting high quality studies with the aim of understanding the relative role of asymptomatic and presymptomatic transmission is instrumental to developing the most informed policies on reopening our cities, states, and countries.

## Introduction

Understanding how SARS-CoV-2 is transmitted is a question that has been at the forefront of efforts to curtail the pandemic. On January 14, 2020, the World Health Organization announced that “there is no clear evidence of human-to-human transmission” of SARS-CoV-2.^[1]^ Six days later, when WHO announced evidence of human to human transmission, countries were left scrambling to enact policy to identify and isolate the ill.^[2]^ Only after these efforts failed, were the more comprehensive quarantine and isolation policies enacted in cities like Wuhan, China. In the absence of definitive evidence of asymptomatic transmission, these intervention policies were made out of an abundance of caution. Understanding the temporal dynamics of SARS-CoV-2 transmissibility is key to safely and successfully reopening our cities, states, and countries until the development of an effective vaccine. Unfortunately, with nearly 8 million confirmed cases and over 434,000 deaths, there is still confusion and a dearth of adequate research around the dynamics of transmissibility of SARS-CoV-2 in the general population.^[3]^ On June 8^th^, 2020, WHO official Maria Van Kerkhove said that asymptomatic transmission of the coronavirus was “very rare.” However, she later clarified this statement saying, “the available evidence from contact tracing reported by Member States [of WHO] suggests that asymptomatically-infected individuals are much less likely to transmit the virus than those who develop symptoms.”^[4]^ Given the absence of definitive information, and because of the importance of this question, there is an urgent need to direct high quality studies towards examining asymptomatic and presymptomatic transmission of SARS-CoV-2.

Asymptomatic individuals are defined as individuals who test RT-PCR positive, but lack symptoms that would indicate SARS-CoV-2 infection. While some individuals may go the entire course of infection and never experience symptoms, other individuals who initially present as asymptomatic may go on to develop symptoms days or weeks later. The individuals who will later develop symptoms are defined as being presymptomatic.

The first large scale reporting of asymptomatic SARS-CoV-2 infection occurred on the Diamond Princess cruise ship, where an estimated 17.9% of cases on board were asymptomatic. ^[5]^ The phenomenon of asymptomatic SARS-CoV-2 infection has since been established in multiple studies, including a UCSF study that found that 53% of individuals who tested positive were not experiencing symptoms at the time of the test.^[6]^ While the existence of asymptomatic cases is well understood, the link between asymptomatic/presymptomatic cases and transmissibility is more tenuous. RT-PCR testing can tell us whether there is detectable virus present, but it does not accurately tell us whether an individual is contagious.^[7]^ Infectivity in cell culture is the standard for determining whether a patient is infectious. In the absence of viral culture data, viral load or cycle threshold (Ct) values derived from RT-PCR data has been used as a proxy for the likelihood of transmission. The Ct is the number of replication cycles required for a signal of RT-PCR product to cross a determined threshold. This value is inversely proportional to the amount of target nucleic acid or viral load in the sample – in particular, high Ct values indicate low viral load. In a study of 90 patients with SARS-CoV-2 infection, Bullard and colleagues found that virus was only successfully isolated when Ct value was below 24.^[8]^

### Asymptomatic viral dynamic studies – current understanding

A small number of studies have attempted to look at viral dynamics in asymptomatic and presymptomatic individuals. One study, from a skilled nursing facility in Kings County, Washington, found viral growth in a patient sample with a cycle threshold (Ct) value of 34, as well as viral growth in asymptomatic and presymptomatic individuals.^[9]^ However, findings in elder care facility may not reflect the general population. It is difficult to recognize early signs and symptoms of respiratory viral infections in elderly populations due to impaired immune responses associated with aging and the high prevalence of preexisting and underlying conditions, such as chronic cough and cognitive impairments. Furthermore, elderly and infirm patients have blunted physiological responses that may allow them to remain apparently asymptomatic during infection. Influenza, another respiratory virus, often manifests with few or atypical symptoms in this population, resulting in confounding of when symptoms are first reported and undermining efforts to isolate ill patients.^[9]^ A second report, looking at individuals exposed during a flight from China to Frankfurt, identified one case of asymptomatic infection and one case of presymptomatic infection with positive culture infectivity.^[10]^ This study does not provide information about the passengers’ health or age, and there is likely to be a bias to downplay mild or moderate symptoms in the context of being detained while traveling. Although these studies have attempted to look at viral dynamics in asymptomatic and presymptomatic individuals in specific populations, to date the authors are not aware of any studies that have successfully cultured live virus from asymptomatic or presymptomatic individuals in the general population.

Despite the absence of live virus isolation and culturing in the general population, many studies and reports have concluded asymptomatic and presymptomatic transmission are prevalent in this pandemic.^[11]^ Modeling studies that are being utilized to predict future case spread and determine the most effective interventions are fundamentally rooted in an understanding of asymptomatic and presymptomatic transmission.

### The basis for asymptomatic and presymptomatic transmission in other viral infections

Viral illnesses have varying transmission profiles. Seasonal influenza is characterized by having peak viral load one day after symptom onset, and individuals generally have detectable levels of RNA from two days before clinical symptoms appear to eight days afterward.^[12]^ Although asymptomatic and presymptomatic individuals may shed influenza virus, studies have not determined if such people effectively transmit influenza.^[13]^

Other viral illnesses like MERS, SARS and Ebola are notable because infectivity appears to increase later in course of illness. MERS-CoV concentrations peaked during the second week of illness.^[14]^ Ebola virus does not appear to have presymptomatic transmission, though individuals can remain infectious for long periods of time after symptoms resolve.^[15]^ Notably, in the case of SARS-CoV, infectiousness peaked 7-10 days after symptom onset.^[16]^

Understanding the viral dynamics and transmission profile of a virus is critically important because it informs the most effective outbreak curtailment strategies. In the case of SARS-CoV and Ebola viruses, efforts aimed at sequestering the ill and contact tracing are highly effective. In the case of influenza virus, contact tracing must extend to the presymptomatic phase, and more aggressive prophylactic containment strategies are necessary. Efforts to curtail SARS-CoV-2 virus will rely on successful contact tracing to halt further transmission. Decisions on how far back to trace contacts and if/when to test asymptomatic contacts will rely on a comprehensive understanding of asymptomatic and presymptomatic transmission.

The aim of this review is to summarize the literature that informs the current understanding of the presence and prevalence of asymptomatic and presymptomatic transmission through studying viral kinetics, case reports, and calculation of serial interval.

## Methods

### Search Strategy

All efforts were taken to comply with PRISMA standards (see Supplement Fig. 1). However, due to the rapidly changing vocabulary and information regarding the SARS-CoV-2 pandemic, this study was not preregistered. Articles for this review were extracted from a PubMed search conducted on June 10, 2020. Articles had to either contain the phrase SARS-CoV-2 or COVID-19 as well as one of the following phrases: presymptomatic transmission, asymptomatic transmission, viral dynamics, viral kinetics, virological analysis, or serial interval. The exact search phrase was: ((“SARS-CoV-2” or “COVID-19”) AND (“presymptomatic transmission” OR “asymptomatic transmission” OR “viral dynamics” OR “viral kinetics” OR “virological analysis” OR “serial interval”)). No year restrictions were applied during search, but due to the fact that SARS-CoV-2 is a novel coronavirus, all studies were from 2020.

### Study Selection

We assessed the eligibility of the studies retrieved during the PubMed search through a two-stage screening process. We first screened the titles and abstracts of all articles. Reviews, correspondence, duplicate references, articles written in languages besides English, and studies that did not mention data collection were excluded.

For all studies that appeared relevant, the full text was reviewed using the same screening procedure delineated above. However, additional filters were added to serial interval studies. Serial interval studies that refit data as well as small studies without supporting statistics, were excluded in this review. Studies excluded during full text analysis are marked ‘initially included then excluded’ in the supplement Table 1.

### Article Types

There were initially 72 results. 2 additional records were added because studies in the review referenced or analyzed their data. After screening, 34 articles met all review criteria (see Fig. 1). These 34 articles fell into the broad categories of (1) case reports, (2) viral dynamic studies, or (3) analysis of serial interval between linked generations of cases. Each of these broad categories will be discussed separately in a subsequent section.

**Figure 1.**
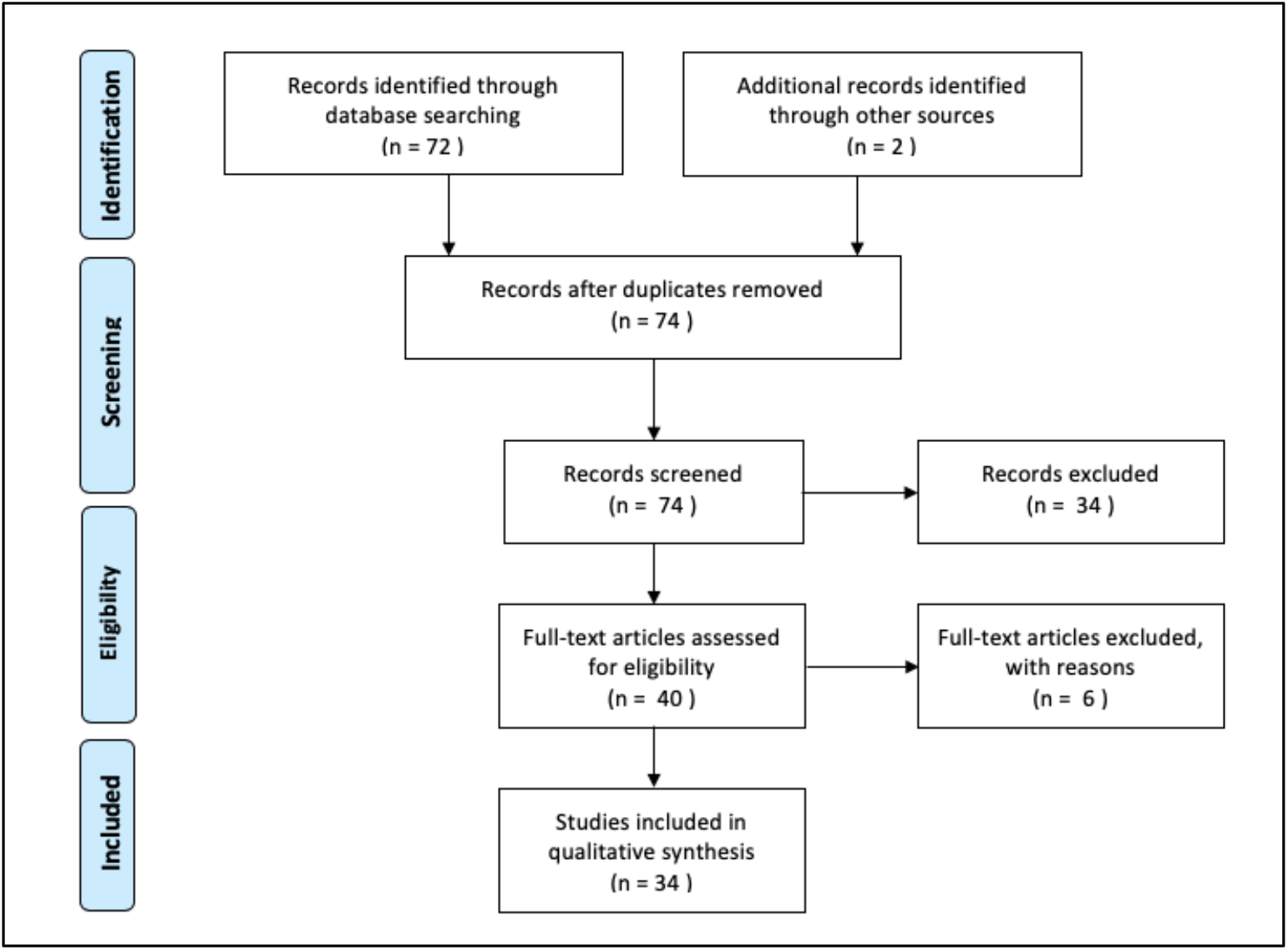
From: Moher D, Liberati A, Tetzlaff J, Altman DG, The PRISMA Group (2009). Preferred Reporting Items for Systematic Reviews and Meta-Analyses: The PRISMA Statement. PLoS Med 6(7): e1000097. doi:10.1371/journal.pmed1000097. A complete list of studies retrieved from the search can be found in the appendix.

### Data Extraction

Case reports and viral dynamic studies were analyzed qualitatively. In case reports, information about the location of the patients, number of patients observed, and notable conclusions were extracted. In viral dynamics studies, number of patients, location, disease severity, testing schedule and sample collection, and notable conclusion were extracted.

For studies that measured serial interval, the mean and/or median serial interval was extracted and compiled along with 95% confidence intervals although other comprehensive methods of expressing data distribution was allowed and noted. Information about the distribution, truncation, and standard deviation was also extracted when available. If study reported serial interval statistics for both high confidence and low or moderate confidence paired transmission statistics, both sets of data were extracted, but the high confidence data was preferentially reported.

Data extracted from serial interval studies (see Supplement Table 2):

- Date Range
- Location
- Number of patient pairs used in determination of serial interval
- Categorization of subsets of data (i.e. high/low confidence)
- Patient relationship (i.e. familial transmission)
- Any other notable features
- Type of distribution (i.e. normal, Weibull, gamma)
- Presence or absence of truncation (i.e. right truncation, not allowing negative values)
- Mean and/or median
- SD of mean/median
- CI on SD
- Type of error reported on mean/median (i.e. 95% CI, quartile, etc)
- Lower bound of error interval
- Upper bound of error interval
- Summary for inclusion in text

Risk of bias and quality of the studies were assessed through author consensus and discussed in the text of the review. Due to the circumstances of the global pandemic, there may be substantial systematic errors in the data being published. The general objective of this paper is to be descriptive, not to draw conclusions about the validity of individual estimates or determine predictive accuracy from the included studies.

## Results

### Evidence of Asymptomatic and Presymptomatic Transmission in SARS-CoV-2

The flow diagram documenting the results of the literature search are shown in Fig 1.

### Case Studies Suggesting Asymptomatic or Presymptomatic Transmission

Case reports providing insight into asymptomatic and presymptomatic transmission are shown in Table 1.

**Table 1.** A summary of case reports from the literature search that yielded insight into the question of asymptomatic and presymptomatic transmission.

| Articles | Type | Description |
| --- | --- | --- |
| Chen *et al.*<sup>[17]</sup> | Case Report | Reported on a family cluster in Hubei province where it appears a parent transmitted SARS-CoV-2 infection to their children while asymptomatic. Mother had traveled to Wuhan before returning home to Xiangyang. |
| Hu *et al.*<sup>[18]</sup> | Case Report | Contact tracing identified 24 asymptomatic COVID-19 infections in Nanjing, Jiangsu Province. One of these asymptomatic cases appears to be a possible source of infection in three relatives, one of which went on to develop severe pneumonia. |
| Huang *et al.*<sup>[19]</sup> | Case Report & Serial Interval Report | One 22-year-old from Wuhan appears to have infected his cousin and six classmates while presymptomatic. |
| Li *et al.*<sup>[20]</sup> | Case Report | Father appears to have infected his daughters, son-in-law, his son-in-law's wardmate, and his wardmate's family while asymptomatic. 2 family clusters of 6 patients stemming from one possible asymptomatic transmitter. |
| Lytras *et al.*<sup>[21]</sup> | Case Report | Noted the relatively high rates of asymptomatic SARS-CoV-2 infection in repatriation flights in flights from Spain, Turkey, and UK. Postulates about the possibility of presymptomatic transmission. |
| Ochiai *et al.*<sup>[22]</sup> | Case Report | 52 obstetric patients were tested before hospital appointments at Keio University Hospital in Tokyo, Japan. 4% were found to be asymptomatic. No cases of asymptomatic transmission documented. |
| Qiu *et al.*<sup>[23]</sup> | Case Report | In 104 cases from Hunan province hospitals, 5 were identified as asymptomatic. Contact tracing suggests two of these cases infected family members. |
| Tong *et al.*<sup>[24]</sup> | Case Report | Reported on two people who were infected with SARS-CoV-2 and their infections appear to have stemmed from contact with a potentially asymptomatic/presymptomatic colleague. These individuals went on to infect other members of their household. However other sources of SARS-CoV-2 infection were not ruled out. |
| Wei *et al.*<sup>[25]</sup> | Case Report | Investigation into 157 locally acquired cases of SARS-CoV-2 infections in Singapore revealed ten cases in 7 family clusters where presymptomatic transmission appears to have occurred. |
| Wong *et al.*<sup>[26]</sup> | Case Report | Identifies two asymptomatic infected individuals from a cluster of cases in the Seri Petaling Mosque in Kuala Lumpur, Malaysia, who appear to have transmitted infection to others. |
| Ye *et al.*<sup>[27]</sup> | Case Report | Family cluster of five patients. One of the patients was believed to be the source of infection, and infected others during a family reunion. Asymptomatic transmission offered as possible explanation. |

The early literature of SARS-CoV-2 asymptomatic transmission was dominated by case reports of apparent asymptomatic transmission, and 9 studies that document cases of apparent asymptomatic or presymptomatic transmission were identified in this systematic review. A majority of these cases were individuals exposed during travel to Wuhan or other cities in Hubei Province, who later transmitted the infection to members of their household or other close contacts.^[17, 19, 20, 23, 24, 27]^ Huang and colleagues reported a cluster of asymptomatic transmission among children, who had rapid onset of illness and various nonspecific or atypical manifestations of illness.^[19]^ While many of these case reports took steps to ensure that those infected by asymptomatic or presymptomatic individuals did not have other plausible sources of infection, they were unable to definitively rule out other sources or community transmission. Other case reports center around regions that were believed to not have community transmission, where exposure to other sources of infection are less likely. One example is the case of a Chinese businesswoman who appeared to have asymptomatically infected some of her colleagues during a work trip in Germany.^[28]^ However, after publication, the supplementary material was modified because the original patient recalled that she was experiencing symptoms during her meetings with colleagues. While this paper did not appear in the keyword search, and is not included in this review, it was frequently cited in other papers analyzed in this review. The subsequent update to the NEJM article is emblematic of the systematic biases in case reports documenting asymptomatic and presymptomatic transmission. Patients or practitioners may make errors when recalling or reporting symptom onset date. Another case report from the keyword search that focuses on areas without broad community transmission reports on seven clusters in Singapore where presymptomatic transmission appeared to be the most likely explanation.^[25]^ This study identified 10 cases where presymptomatic transmission appeared to occur 1-3 days before symptom onset in the initial patient. While compelling, the retrospective nature of these studies makes it difficult to rule out mild symptoms being present during transmission, or other sources of infection.

All case reports of asymptomatic and presymptomatic transmission are confounded by the highly subjective nature of reporting symptom onset and exposure date. Factors like age, cultural norms, and public communication about the pandemic may influence when people report their symptoms beginning. For example, an older person with chronic illness may attribute muscle and joint pain to age, whereas a younger person may call that a symptom. Additionally, as the pandemic has progressed, our categorization of what is considered a symptom has expanded. In February, the WHO said symptoms of COVID-19 included fever, dry cough, fatigue, sputum production, shortness of breath, sore throat, headache, myalgia or arthralgia, chills, nausea or vomiting, nasal congestion, diarrhea, hemoptysis, and conjunctival congestion.^[29]^ In late February, Mao and colleagues first reported that anosmia, or loss of sense of smell, were symptoms of COVID-19, and this finding was supported in additional research.^[30]^ On April 17^th^, the WHO added loss of smell or taste as well as rash and skin discolorations of fingers and toes as additional symptoms of COVID-19.^[31]^ Knowledge of these changing definitions, differing levels of chronic illness, and varying levels of symptom awareness will alter when individuals first report experiencing symptoms.

Two additional reports included in this keyword search inferred the possibility of asymptomatic transmission from positive RT-PCR tests in asymptomatic and presymptomatic individuals. Lytras and colleagues noted a high prevalence of SARS-CoV-2 infection in asymptomatic cases in repatriation flights to Greece.^[21]^ While this study supports the well-documented phenomenon of asymptomatic cases, the possibility of asymptomatic transmission is a hypothetical, as a positive RT-PCR test does not confirm that an individual is contagious. This study failed to provide insight into the feasibility of actual transmission during presymptomatic or asymptomatic infection because the authors failed to report Ct values of RT-PCR positive individuals, did not culture virus, and did not identify possible transmission chains. The study by Ochiai and colleagues had similar findings and limitations.

### Viral Dynamics

Results from the literature search that documented viral dynamics are shown in Table 2.

**Table 2.** Results of literature search that yielded insight into the question of presymptomatic transmission that pertained to the study of viral dynamics. Two of the included studies did not appear in the PubMed literature search and were added later because they appeared as references in other reviewed studies (marked as externally sourced).

| Paper | Included/Excluded in Review | Category | Description |
| --- | --- | --- | --- |
| Ding *et al.*<sup>[32]</sup> | Included | Viral Dynamics | 64 patients from Ruian People's Hospital in Zhejiang, China were retroactively enrolled in this study. Patients received lopinavir/ritonavir, interferon-α regimen, and some also received arbidol. Samples were taken at baseline and then every 2-3 days until discharge. Viral loads peak at start of observation. |
| He *et al.*<sup>[33]</sup> | Included | Viral Dynamics & Serial Interval | 94 patients admitted to Guangzhou Eighth People's Hospital were studied. 414 throat swabs were collected between symptom onset up to day 32. At the time of this review, no information was found about frequency or duration of swab collection. Patients received treatment that is standard of care, including combinations of antivirals, antibiotics, corticosteroids, immunomodulatory agents and Chinese medicine preparations. Viral loads peak at start of observation. |
| Kim *et al.*<sup>[34]</sup> | Included | Viral Dynamics | 10 asymptomatic and 3 presymptomatic individuals were studied. Viral load peaks at the beginning of observation. Patients were observed at Affiliated Hospitals of Chonnam National University between February 4 and April 7, 2020. At the time of this review, no information was found about the frequency or duration of swab collection. Viral loads peak at start of observation. |
| Liu *et al.*<sup>[35]</sup> | Included | Viral Dynamics | 76 patients admitted to the First Affiliated Hospital of Nanchang University (Nanchang, China) from Jan 21 to Feb 4, 2020. 46 cases were mild and 30 were severe. At the time of this review, no information was found about frequency or duration of sample collection or treatment patients were receiving. Found that patients with severe COVID-19 tend to have a high viral load and a long virus-shedding period. Cite previous work showing viral loads peak during first week of disease onset. |
| Lui *et al.*<sup>[36]</sup> | Included | Viral Dynamics | A study of the first 11 laboratory-confirmed COVID-19 patients hospitalized in 2 hospitals in Hong Kong in February 2020. 6 had moderate/mild disease, 5 had severe/critical disease. Authors conclude that viral load appears to peak in the first week in mild cases, and potentially peak later in severe cases. All patients were taking antivirals including lopinavir/ritonavir, ribavirin, beta-interferon, and one patient was taking corticosteroid therapy. |
| To *et al.*<sup>[37]</sup> | Included, externally sourced | Viral Dynamics | 23 patients from 2 hospitals in Hong Kong with laboratory confirmed COVID were entered in this cohort study. Patients were screened between Jan 22-Feb 12, 2020. Ten patients had severe COVID-19, 13 had mild disease. The median interval between symptom onset and hospitalization was 4 days. Five were admitted to ICU and 2 died. All patients produced an early morning saliva sample from the posterior oropharynx. Saliva viral load was also measured. In Fig 2. viral loads appear to peak a few days after symptom onset, but authors concluded viral loads peak around symptom onset. |
| Wölfel *et al.*<sup>[38]</sup> | Included | Viral Dynamics | Viral dynamics determined from 9 individuals from a single cluster in a single hospital in Munich, Germany. All patients were admitted after symptom onset. For most of the patients, viral loads appear to peak around the time observation began. At the time of this review, no information was found on patient treatments. |
| Yoon *et al.*<sup>[39]</sup> | Included | Viral Dynamics | Viral dynamics in diverse body fluids of 2 patients were studied. Patients were sampled every 2 days on hospital days 1-9. Patient 1 received lopinavir/ritonavir 400/100mg twice a day along with hydroxychloroquine 400 mg once daily. Patient 2 received lopinavir/ritonavir 400/100mg twice a day. |
| Young *et al.*<sup>[40]</sup> | Included, externally sourced | Viral Dynamics | Studied first 18 patients diagnosed with SARS-CoV-2 infection in Singapore between January 23 and February 3, 2020. 5 received lopinavir-ritonavir. For half of patients presented at hospital more than 2 days after symptom onset. Viral loads in nasopharyngeal samples from patients with COVID-19 peaked within the first few days of observation before declining. |
| Zhou *et al.*<sup>[41]</sup> | Included | Viral Dynamics | This study included 31 adults with confirmed SARS-CoV-2 infection who were asymptomatic on admission. No information about patient treatment. 22 of the patients went on to develop symptoms while 9 remained asymptomatic. When comparing the viral dynamics of asymptomatic and presymptomatic individuals, Zhou *et al.* found asymptomatic individuals had lower Ct values, and had peak viral loads in the second week of hospitalization. |

Studying temporal viral dynamics allows for the prediction of peak infectiousness. In this review there were ten studies that measured viral temporal dynamics and kinetics of SARS-CoV-2. Eight of these studies measured viral dynamics by quantifying successive nasopharyngeal swabs in hospitalized patients. The two remaining papers focused exclusively on asymptomatic and presymptomatic individuals. From the eight studies of viral dynamics in hospitalized patients, all patients except one in the Zou *et al*. paper were symptomatic. The one asymptomatic individual in Zou *et al*. remained asymptomatic throughout the course of the study.

The eight studies reported viral loads were at their highest levels around the time observation began. Therefore, the authors of these studies concluded viral loads peak close to when symptoms emerge. However, this discovery must be prefaced by the limitation that all patients in the studies were enrolled after symptom onset, and therefore presymptomatic viral loads were not measured. This shortcoming is further propagated by the fact that patients often will not see a clinician immediately after symptom onset, in these cases we cannot rule out the possibility that viral load peaks after symptom onset. While studying COVID-19 in China, Zhang and colleagues found that an average of 2.5 days elapsed between symptom onset and first healthcare consultation.^[56]^ Although this decreased from 3.0 to 1.6 days as the pandemic progressed. If individuals are only infectious for 8 days, as Bullard and colleagues report, this delay in seeking care greatly confounds our ability to measure comprehensive viral dynamics.^[8,38]^ Additionally, the studies do not disclose how soon the first swab was taken after symptoms were reported; a margin of error of a day might dramatically change the viral load in patients. While the finding that viral load appears to peak soon after symptoms are detected in patients suggests that presymptomatic transmission is plausible, there is not enough information about the distribution of SARS-CoV-2 viral kinetics in presymptomatic stage to conclude when infectiousness begins.

When modeling viral dynamics, basic assumptions about the distribution will have dramatic effects on our prediction of when infectivity begins, and the specific time between symptom onset and viral load tests can dramatically change our understanding of transmissibility and infectiousness. Examples of hypothetical distributions of SARS-CoV-2 viral load and their effect of predicting transmissibility are shown in Fig 2. Knowledge of the shape of the distribution will impact our responses to curtail the pandemic.

**Figure 2.**
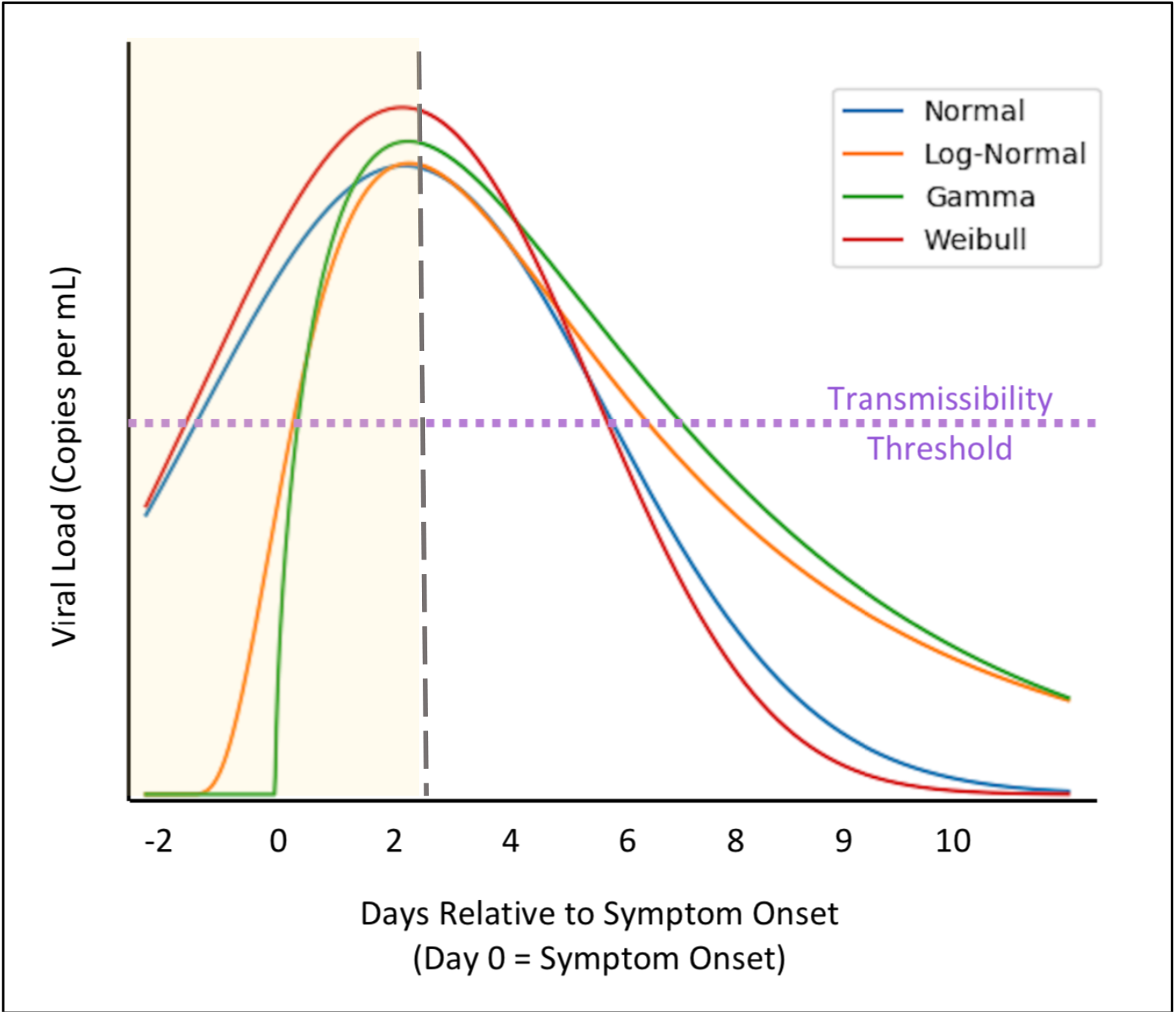
Hypothetical distributions of SARS-CoV-2 viral load. Different assumptions about the shape of the distributions will impact when and if presymptomatic transmission will occur. A line indicating the threshold of transmissibility is shown in purple, which is currently believed to be 10^6^ copies per mL. The intersection of the purple line with the various curves would show when an individual becomes contagious. In these hypothetical distributions, a normal and Weibull distribution suggest significant presymptomatic transmission, while a gamma and lognormal distribution seem to suggest limited presymptomatic transmission. These conclusions can change with different transmission thresholds and distribution parameters. A vertical dashed line in grey shows when an individual might seek medical consultation, which Zhang and colleagues report as being 2.5 days after symptom onset in China during the COVID-19 pandemic.^[53]^ Although this number decreased from 3.0 to 1.6 days as the pandemic progressed. Assuming patients don’t seek medical care for 2.5 days, the light-yellow shaded region refers to the area where data is lacking. While many studies concluded viral load peaks when observation begins, for almost all of the studies, a significant portion of time elapsed between when symptoms first appeared and observation began.

Wolfel and colleagues attempted to relate RT-PCR quantification of viral load with infectivity. The authors combined RT-PCR measurement with viral culturing and found that the success of virus isolation in culture was a function of viral load: only samples that contained greater than 10^6^ copies per mL yielded an isolate (although Ct value was not reported in this study, He *et al*. reports this corresponds to a Ct value of 24).^[42]^ Interestingly no isolates were obtained after day 8, despite continuing high viral loads. This finding suggests persistent RNA detection represents non-viable virus that is not infectious. This finding demonstrates that while viral load can be predictive of transmissibility, it is not a perfect correlation. The viral studies of Wolfel *et al*., Lui *et al*., To *et al*., Young *et. al*. and Yoon *et al*. were limited by small sample size. However, He *et al*., Liu *et al*., and *Ding et al*. have similar findings with larger sample sizes.

Despite the attempt to comprehensively profile SARS-CoV-2 kinetics, all eight of these studies were limited in their scope because they were not able to swab patients before symptom onset. An additional limitation of these studies is that many failed to specify the exact schedule of when patient swabs were collected. Only one study, To *et al*. mentioned a precise collection schedule that applied to all patients. It is also worth noting that nasopharyngeal swabs are an imperfect proxy for viral production. Studies on influenza have shown variability in viral load when sampling left and right nostrils and this finding will likely be similar for SARS-CoV-2.^[43]^ Perhaps the most important limitation of these studies is that the studies either did not specify or did not exclude individuals who were undergoing treatment. Undergoing antiviral, interferon, or steroid therapy may disrupt the natural progression of viral load. While the study by Ding and colleagues had the purpose of examining the viral kinetics during antiviral treatment, data focusing on viral load after therapeutic interventions cannot provide insight into the viral dynamics of the natural history of infection. Antiviral and interferon treatments should diminish viral replication and artificially cause viral load to peak at the start of treatment, while steroid treatment may dampen the immune response and potentially cause viral replication to increase. If the viral load data is a basis for clinical decision making, this will even further confound results because an increasing viral load would be the basis for more extensive interventions and therapeutic treatment.

There is an urgent need to study the viral kinetics in presymptomatic individuals. Kim *et al*. analyzed the Ct values of three presymptomatic patients and found the highest levels of virus were one to two days before symptom onset. However, this dataset is extremely small (n=3), and one of the patients was on the threshold of detection. It is hard to reliably extract general trends from this limited sample. Zhou *et al*. studied the viral dynamics of 31 patients who were asymptomatic upon hospital admission for laboratory confirmed SARS-CoV-2 infection. Twenty-two of the patients went on to develop symptoms while nine remained asymptomatic by the case definition used in the study. When comparing the viral dynamics of asymptomatic and presymptomatic individuals, Zhou *et al*. found asymptomatic individuals had lower Ct values, and had peak viral loads in the second week of hospitalization. This data cannot be extrapolated to inform our understanding of presymptomatic viral dynamics because symptom onset date was not disclosed, and therefore the viral load data cannot be ascertained in relation to symptom onset.

While there currently appears to be consensus that viral load appears to peak with the beginning of observation, these studies are preliminary and that there is a dearth of data regarding infectiousness during the presymptomatic interval. In evaluating viral dynamics, knowledge of the shape of the distribution would be valuable to our understanding of transmissibility of SARS-CoV-2.

### Serial Interval Between Generations of Cases

Another approach to uncovering the prevalence of presymptomatic transmission has relied on calculations of serial interval. Serial interval is defined as the time between symptom onset in the first-generation case and the second-generation case. This method requires identification of serial cases where one individual (first-generation case) infected another individual (second-generation case). If the observed mean serial interval is shorter than the incubation period, this would support the conclusion that a significant portion of transmission may have occurred presymptomatically.

Fourteen papers in this review calculated serial interval by looking at paired cases with probable point transmission linkage (see Table 3).

**Table 3.** Results of literature search that yielded insight into the question of presymptomatic transmission that pertained to serial interval. Studies that were excluded after full text analysis were also included.

| Paper | Included/Excluded in Review | Category | Description |
| --- | --- | --- | --- |
| Aghaali *et al.*<sup>[44]</sup> | Included | Serial Interval | Study calculated serial interval for 37 linked cases in Qom, Iran, who were identified through contact tracing. Due to limited availability of RT-PCR tests, second generation cases were confirmed with chest CT. Authors assumed a gamma distribution of serial intervals. |
| Bi *et al.*<sup>[45]</sup> | Included | Serial Interval | Serial interval calculated from 48 pairs with clear relationship between index case and secondary case. Data released by Shenzhen CDC. A gamma distribution of serial interval times was used. |
| Böhmer *et al.*<sup>[46]</sup> | Included | Serial Interval | Data sourced from one outbreak cluster and their contacts in Germany. Altogether 16 paired transmission events were reported. At the time of this review, no distribution was reported. |
| Du *et al.* MedRxiv<sup>[47]</sup> | Included | Serial Interval | 339 confirmed cases of COVID-19 identified from 264 cities in mainland China prior to February 19, 2020. Sourced public data. Authors looked at multiple distributions for serial interval, but ultimately chose a normal distribution. The authors found household transmission led to shorter serial interval than non-household transmission inside the household (4.57 days [95% CI 3.76–5.38]) versus outside the household (5.85 days [95% CI 5.06–6.64]). |
| Du *et al.*<sup>[48]</sup> | Included | Serial Interval | Identified 468 paired cases from provinces outside of Hubei Province in China. Similar to analysis listed above, which appears to be an earlier version of this study. Authors assumed a normal distribution of serial intervals. (The authors ruled out gamma or Weibull distribution). |
| Ganyani *et al.*<sup>[49]</sup> | Included | Serial Interval | Studied 54 cases in Singapore and 114 paired cases in Tianjin, China that were part of outbreak clusters. Authors included cases in clusters with likely but not definitive transmission links. Authors determined density function of serial intervals by using a Monte Carlo estimation. They then used bootstrap sampling to determine confidence intervals. |
| He *et al.*<sup>[33]</sup> | Included | Serial Interval and Viral Dynamics | 77 transmission pairs were sourced from publicly available information from multiple countries. Data was fitted to a gamma distribution of serial intervals. |
| Kwok *et al.*<sup>[50]</sup> | Included | Serial Interval | Serial intervals were estimated from 26 (probable: 9; certain: 17) paired data from Hong Kong Centre for Health Protection (CHP) before February 13, 2020. Authors used a lognormal distribution of serial intervals, but gamma and Weibull distributions were also examined. |
| Nishiura *et al.*<sup>[51]</sup> | Included | Serial Interval | Identified 28 paired cases, 18 of which were considered high quality. The data was fit to many different distributions, but authors ultimately chose Weibull distribution of serial intervals as best fit for high quality data. Data sourced from articles and government documents. |
| Wang K. *et al.*<sup>[52]</sup> | Included | Serial Interval | 27 cases with transmission chains were identified and studied in Shenzhen, China. Transmission events sourced from publicly released information and identified 27 transmission chains, including 23 infectees matched with only one infector. Authors used a Weibull distribution of serial intervals (but also looked at other distributions). |
| Wang X. *et al.*<sup>[53]</sup> | Included | Serial interval | Enrolled 37 cases and found 9 transmission chains. From these 9 paired cases, the authors calculated serial interval and assumed gamma distribution of serial intervals. Patients were seen at Wuhan Union Hospital between January 5 to February 12, 2020. |
| Wu *et al.*<sup>[54]</sup> | Included | Serial Interval | Studied 48 secondary cases stemming from household transmission. Fit to lognormal distribution. Zhuhai, China. Enrolled index cases and studied their household members. |
| You *et al.*<sup>[55]</sup> | Included | Serial interval | Data sourced from 198 linked transmission cases outside Hubei Province as of March 31, 2020. No information was found on the type of distribution used, and statistics were reported as interquartile range. |
| Zhang *et al.*<sup>[56]</sup> | Included | Serial Interval | Serial Interval calculated from 35 secondary cases stemming from 28 primary cases. Serial interval was fit to a gamma distribution of serial intervals (although other distributions were analyzed as well). Data taken from provinces outside Hubei. |
| Huang *et al.*<sup>[19]</sup> | Initially included, then excluded from Serial Interval Data | Serial Interval and Case Report | Data about serial interval excluded because general population was not studied. Study focused exclusively on young individuals. |
| Li *et al.*<sup>[57]</sup> | Initially included then excluded | Serial Interval | This study used prior assumptions from SARS-CoV data in their calculation of serial interval, therefore study was excluded. |
| Pung *et al.*<sup>[58]</sup> | Initially included then excluded | Serial Interval | Study was of the first three clusters in Singapore, which identified 3 paired transmission cases. Study was excluded because no statistics on data were provided, and primary data could not be located. |
| Son *et al.*<sup>[59]</sup> | Initially included but then excluded | Serial Interval | Study of patients in Busan. Authors report mean serial interval as 5.54 days [95% CI 4.08-7.01 days]. Excluded because full article was not available in English. |

Most of these fourteen reports in Table 3 calculated serial interval by compiling data from publicly available sources or from municipal datasets. It is difficult to control for quality and bias from these publicly available reports. These datasets are compiled from human-to-human transmission reports from different countries, jurisdictions, and points in time. These factors may impact standards of reporting cases or symptom onset. In addition to bias or error in the publicly sourced data, all of the serial interval studies are confounded by their reliance on self-reported symptom start date. As stated earlier in this paper, what is considered a symptom varies by region, culture, age, and time, and the definition of symptoms has become more expansive as time has progressed. For example, patients who notice loss of smell may have an earlier symptom start date than a patient who only reports fever and dry cough. The date reported as the onset of symptoms is also subject to error due to inherent inaccuracy of memory. Furthermore, in the datasets, the authors report the date of symptom onset rounded to the nearest day. This is especially problematic because the difference in serial interval and incubation period calculated in these studies often differed by less than a day. It is therefore not possible to ascertain if the difference between calculated serial interval and incubation period are true differences, or an artefact of rounding error.

Many studies of serial interval are biased towards household transmission because it is more straightforward to isolate transmission chains and rule out other sources of infection in a household setting. In household transmission cases, newly infected individuals will likely be exposed to a much higher dose of viral particulates than would occur in a more casual transmission case. Exposure to higher inoculum may result in a decreased incubation period for household transmission. Given that the papers compared serial intervals to estimates of incubation period, the difference in inoculum between household transmission and community transmission may account for the difference between the calculated serial interval and incubation period. The interpretation of this data is further complicated by the fact that estimates of incubation period vary between studies.

Mean serial interval was preferentially reported for the studies in this review and is denoted as a red circle on the graph of Figure 3. When mean was not reported, median was used (denoted with a red triangle). It is important to note that in skewed distributions such as gamma and lognormal, median is often less than mean. Despite the various possible sources of error and bias, it is notable that almost all of the studies have calculated serial intervals that fall within the 95% CI of the estimated incubation period as reported by Li and colleagues.^[58]^ This finding is compatible with the hypothesis that infectiousness appears to emerge at symptom onset. This interpretation is qualitative and should be revisited through meta-analysis and further study.

**Figure 3.**
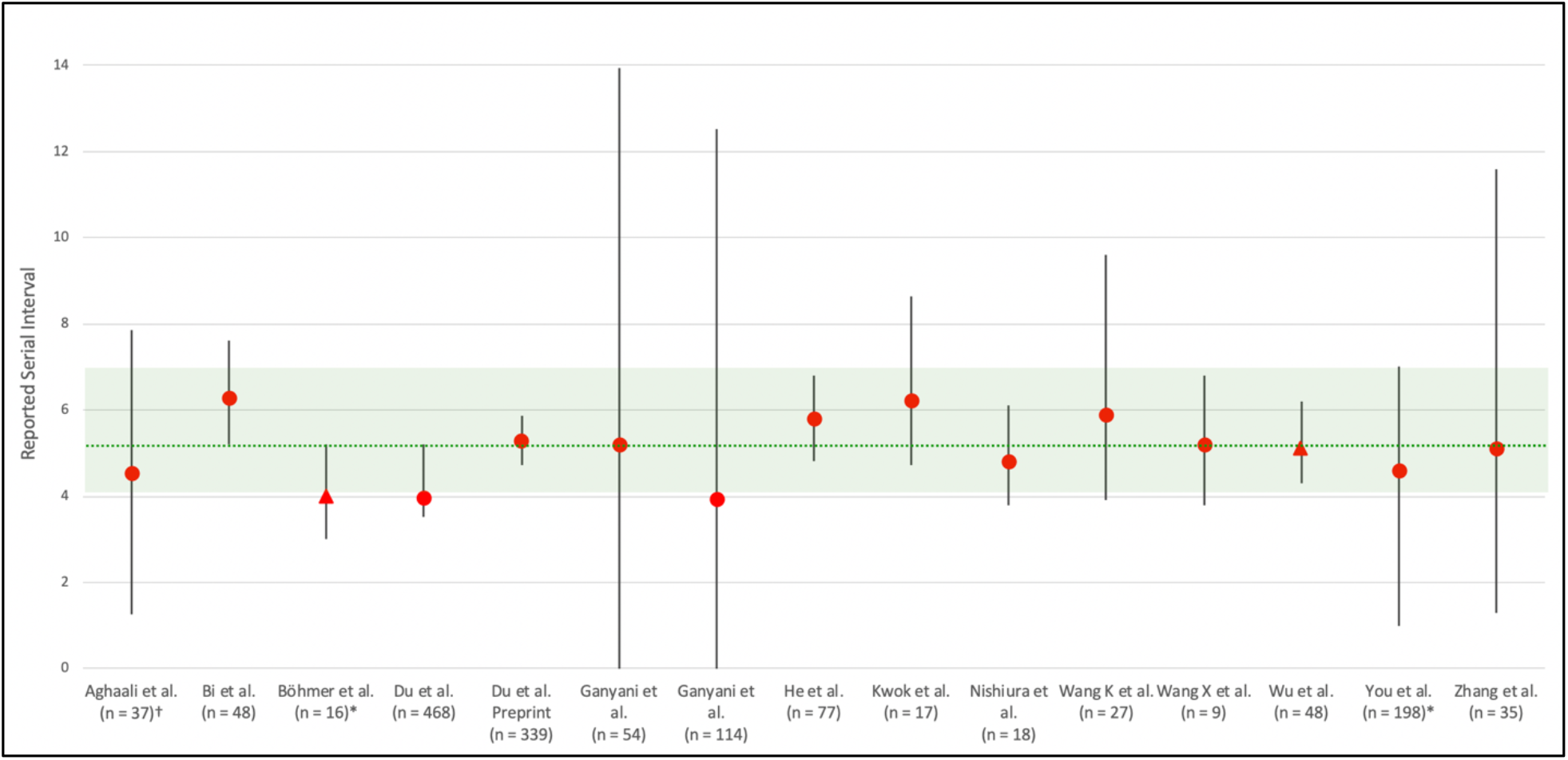
Green dotted line shows the reported mean incubation period of 5.2 days. Green shaded area shows 95% CI of incubation period as reported by Li *et al*. We preferentially reported the mean serial interval (red circle). If mean was not reported, median was used (red triangle). However, it should be noted that in skewed distributions such as gamma and lognormal, median is often less than mean. In the case of Wu *et al*. the mean was noted as 6.3, but no error terms were reported, therefore median was used in the figure. Error bars default to show 95% CI on serial interval on statistic, however if 95% CI was not reported, 1st and 3rd quartiles were used (denoted by *) or +/- 1 standard deviation (denoted by †). Error bars that extended below zero were not shown but are reported in supplemental Table 2. The two studies from Du *et al*. may use overlapping data, and if so, these serial intervals cannot be considered independently.

## Discussion

This review focused on primary publications that reported asymptomatic and presymptomatic transmission through case reports, viral kinetics studies, and serial interval calculations. These different approaches have methodological shortcomings, which are summarized in Table 4.

**Table 4.** Summary of the major sources of error observed in the literature that was reviewed in this analysis.

| Shortcoming | Description | Type of Study Most Affected |
| --- | --- | --- |
| Studying non representative populations | Age, chronic illness, and other factors can impact perception and reporting of symptoms. For example, it is difficult to recognize early signs and symptoms of respiratory viral infections in elderly populations, due to impaired immune responses associated with aging and the high prevalence of preexisting and underlying conditions, such as chronic cough and cognitive impairments. Furthermore, elderly and infirm patients have blunted physiological responses that may allow them to remain asymptomatic during infection. On the other hand, younger individuals may be more likely to remain asymptomatic. Studies can only represent the demographic they study. | All studies |
| Small sample size | Small sample sizes are more subject to bias and skewed results. | All studies |
| Errors when recalling or reporting symptom onset date | Many studies of transmission and serial interval recall on patients self-reported symptom onset date. Recall bias and other errors can alter when an individual reports symptom onset date. | Case reports and serial interval |
| Errors in determining sources of infection | In case report and serial interval studies it is impossible to rule out other sources of infection. This confounds determining if presymptomatic transmission occurred. Future studies can use viral genome sequence to better determine source of infection. | Case reports and serial interval |
| Varying definition of symptoms | What is considered a symptom varies by region, culture, age, and time. In February, symptoms of COVID-19 included fever, dry cough, fatigue, sputum production, shortness of breath, sore throat, headache, myalgia or arthralgia, chills, nausea or vomiting, nasal congestion, diarrhea, hemoptysis, and conjunctival congestion. In April, the WHO added loss of smell or taste as well as rash and skin discolorations of fingers and toes as additional symptoms of COVID-19. | Case reports, and serial interval |
| Household transmission altering incubation period | In household transmission cases, newly infected individuals will likely be exposed to a much higher dose of viral particulates than would occur in a more casual transmission case. Exposure to higher inoculum may result in a decreased incubation period for household transmission. | Serial interval |
| Effect of treatment on viral kinetics | Undergoing antiviral, interferon, or steroid therapy may disrupt the natural progression of viral load. Antiviral and interferon treatments should diminish viral replication and artificially cause viral load to peak at the start of treatment, while steroid treatment may dampen the immune response and potentially cause viral replication to increase. If the viral load data is a basis for clinical decision making, this will even further confound results because an increasing viral load would be the basis for more extensive interventions and therapeutic treatment. | Viral Dynamics |
| Using RT-PCR test as a proxy for infectiousness | RT-PCR testing informs clinicians whether there is detectable virus present, but it cannot determine whether an individual is contagious. Infectivity in cell culture is the standard for determining whether a patient is infectious, but even this is a proxy for transmissibility. Currently, it is believed a Ct value below 24 is the threshold for being infectious. | All studies |
| Inferences about viral load distribution before samples collected. | The finding that viral load is highest around the time symptoms are detected in patients suggests presymptomatic transmission is plausible. However, there is not enough information about the distribution of SARS-CoV-2 viral kinetics in presymptomatic stage to infer when infectiousness begins. Basic assumptions about the distribution will have dramatic effects on our prediction of when infectivity begins, and the specific time between symptom onset and viral load tests can dramatically change our understanding of transmissibility and infectiousness. | Viral dynamics, and serial interval |
| Not measuring viral load in presymptomatic stage | Viral loads generally appeared at their highest levels when observation in the clinical setting began. Therefore, authors have concluded viral loads peak when symptoms emerge. However, Zhang *et al.* has shown that multiple days elapse between symptom onset and seeking clinical care. This makes it even more difficult to extrapolate viral peak and presymptomatic viral dynamics. Without measuring viral load in the presymptomatic phase, the dynamics during the presymptomatic period can only be hypothesized. | Viral Dynamics |
| Rounding errors during calculation of incubation period and serial intervals | The datasets from the papers in this review that measured serial interval rounded the date of symptom onset to the nearest day. This is problematic because the difference in serial interval and incubation period calculated in these studies often differed by less than a day. It is therefore difficult to know if the difference between calculated serial interval and incubation period are true differences, or an artefact of rounding error. | Viral dynamics, and serial interval |
| Sampling errors in nasopharyngeal swabs | Nasopharyngeal swabs are an imperfect proxy for viral production. Studies on influenza have shown variability in viral load when sampling left and right nostrils and similar findings will be found in SARS-CoV-2. Any study on viral dynamics must account for high levels of variability in swab samples. | Viral Dynamics |

### Case Studies

While case studies, in aggregate, can offer compelling insight into the existence of asymptomatic and presymptomatic transmission, these reports have many limitations. Even if broad community transmission is not observed, it is still extremely difficult to rule out other sources of infection. Future studies should use viral sequence to more precisely determine sources of infection and transmission chains. Additionally, the temporal variation in what is classified as a symptom of COVID-19, combined with bias and reporting errors, make anecdotal reports of symptom start date unreliable. These factors confound the case reports that highlight asymptomatic or presymptomatic transmission and make it difficult to draw reliable conclusions.

### Viral Dynamics

The preliminary SARS-CoV-2 viral dynamics studies demonstrate that viral titer peaks at patient presentation. Without more knowledge of the temporal distribution of viral load, presymptomatic transmission cannot be conclusively shown. In interpreting viral dynamic studies, a sharp rise in viral load, as would be observed if viral load followed a lognormal or gamma distribution, may link infectiousness with the start of symptom onset. On the other hand, a normal or Weibull distribution in viral load supports the possibility of presymptomatic transmission. It is important that the viral dynamics data be validated with culture data on infectivity. As Wolfel and colleagues demonstrated, while viral load is a proxy for infectivity and transmissibility, it is not perfectly correlated.

Nasopharyngeal swabs are an imperfect proxy for viral production, and any study on viral dynamics must account for high levels of variability in swab samples. Future research efforts should focus on other methods of virus harvesting including throat, blood, fecal, or urine samples, and must prioritize quantifying viral load from individuals in the presymptomatic stage.

### Serial Interval

The shape of the distribution has the most direct impact on studies attempting to measure serial interval between successive generation of cases. Differences in the assumption about the distribution of the viral load curves can alter the calculation of how much presymptomatic transmission is occurring. Articles measuring serial interval in this review assumed Weibull, gamma, lognormal, and normal distributions. Furthermore, serial interval calculations in the reported literature rely heavily on cases of household transmission. It is not possible to differentiate an observation of shortened serial interval due to presymptomatic transmission from a decreased incubation period due to higher inoculum in household transmission.

This systematic review attempted to comprehensively document and analyze literature on asymptomatic and presymptomatic transmission of SARS-CoV-2. It is worth noting, especially because of the rapidly evolving nature of the COVID-19 pandemic, there is likely a risk of systemic bias in manuscripts published on this topic. Although all efforts were made to include a comprehensive review of the literature, the rapid progression and influx of new publications, as well as prevalence of preprint manuscripts on this topic, mean this literature review was likely affected by incomplete retrieval of identified research.

### Proposed study to characterize presymptomatic transmission

In order to ascertain the temporal viral dynamics and transmissibility of SARS-CoV-2, it is important to study a representative healthy population before, during, and after SARS-CoV-2 infection. It is essential to combine RT-PCR data with viral culturing data to ascertain transmissibility. In particular, such a study would clarify when viral load and transmissibility commence relative to the time of infection, and peak relative to the onset of symptoms. As well as provide insight into the relationship between viral load or Ct and the severity of symptoms. Additionally, this type of study would be instrumental in determining the most appropriate distribution curve to characterize the rise and decline of viral infectivity.

This study needs to involve a sufficient number of volunteers tested at frequent intervals to obtain a clear answer. Samples need to be collected in a consistent manner using the most reliable available tests. Because of the logistics of such a study, it would be valuable to collect additional information regarding subject demographic features as well as biochemical, immunological, and genetic markers that may be predictive of viral dynamics and transmissibility. Of particular interest will be the impact of age on viral load and infectivity. Among infected individuals, the additional determination of viral genomic sequences would allow for molecular epidemiological analysis of transmission between specific individuals as well as determine any differences in viral load profiles due to mutations in the virus.

One way to accelerate the determination of viral kinetics is to focus on a population with high risk of infection and low risk of complications, such as workers in factory at the start of an outbreak, or individuals identified through contact tracing. This population would be ideal to study because these individuals would likely not seek treatment for SARS-CoV-2 infection, therefore the viral dynamics data would not be confounded by therapeutic interventions like antiviral therapy.

While many of the research studies highlighted in this review have supported asymptomatic and presymptomatic transmission, these studies have been inadequate to ascertain the contribution of asymptomatic and presymptomatic transmission in the spread of SARS-CoV-2 infection. Understanding the temporal dynamics of SARS-CoV-2 transmission from asymptomatic and presymptomatic individuals is critical to our efforts to formulate effective and efficient policies to curtail the pandemic and to minimize the risks associated with phased reopening.

## Data Availability

All data in this manuscript is publicly available.

## Acknowledgments

A special thanks to Dr. Reuben Granich, Dr. Wendy Max, and Jessie Yeung for useful discussion, feedback, and support on this manuscript.

